# The role of connectivity on COVID-19 preventive approaches

**DOI:** 10.1101/2021.03.11.21253348

**Authors:** V. Miró Pina, J. Nava-Trejo, A. Tóbiás, E. Nzabarushimana, A. González-Casanova, I. González-Casanova

**Author notes:** Both contributed equally and should be considered first authors (V. Miró Pina, J. Nava-Trejo). Both should be considered senior authors (A. González-Casanova, I. González-Casanova).

## Abstract

Preventive and modelling approaches to address the COVID-19 pandemic have been primarily based on the age or occupation, and often disregard the importance of heterogeneity in population contact structure and individual connectivity. To address this gap, we developed models based on Erdős-Rényi and a power law degree distribution that first incorporate the role of heterogeneity and connectivity and then can be expanded to make assumptions about demographic characteristics. Results demonstrate that variations in the number of connections of individuals within a population modify the impact of public health interventions such as lockdown or vaccination approaches. We conclude that the most effective strategy will vary depending on the underlying contact structure of individuals within a population and on timing of the interventions.

**Author summary:** The best strategy for public health interventions, such as lockdown or vaccination, depends on the contact structure of the population and the timing of the intervention. In general, for heterogeneous contact structures that mimic the COVID-19 spread, which is characterized by the presence of super spreaders, vaccinating the most connected individuals first was the most effective strategy to prevent infections and deaths, especially when coupled to serological tests. Models considering heterogeneity in human interactions need be used to identify the best potential vaccine prioritization strategies.

## Introduction

The Severe Acute Respiratory Syndrome Coronavirus 2 (SARS-CoV-2), responsible for the coronavirus infectious disease COVID-19, has infected over 165 million and caused more than 3.4 million deaths globally. [1, 2] It continues to spread in most countries despite the wide range of preventive approaches that have been deployed.

In the absence of effective vaccines, country-level responses ranged from strict and prolonged lockdowns aimed at completely stopping transmission, to allowing a substantial proportion of the population to get infected to ultimately reach natural herd immunity, which is when the proportion of individuals who have been exposed to a disease and developed immunity is enough to protect remaining susceptible individuals from exposure and stop the spread of a disease. [3] Even with new evidence suggesting that heterogeneity in age and social connectivity reduces the proportion of individuals that need to be infected to reach herd immunity, the human cost of this approach is unacceptable from a public health perspective, and effective immunization of the population remains the only viable approach to reach herd immunity. [4] Now that effective vaccines against SARS-CoV-2 have been approved, it is imperative to identify prioritization strategies that maximize the impact of the limited doses available to reach herd immunity with the minimum loss of human life. [5]

Most countries have laid out vaccination strategies that prioritize healthcare workers, followed by those living in elderly care facilities, or essential workers (e.g., teachers, food industry), and then age groups from older to younger. [5-8] This prioritization of older populations is based on disease severity and higher risk of death of COVID-19 among this group. However, another popular prioritization strategy - that has been shown to be effective for influenza - is to begin with younger individuals to create an immune shelter around those most vulnerable. [9] Beubar et al. evaluated these two strategies in different contexts and demonstrated that, in most scenarios, years of life lost were minimized by strategies that prioritized adults over 60 years, even as incidence was minimized when younger adults were prioritized. The exceptions to that rule were under low community transmission (basic reproductive number - R_0_ = 1.15 vs 1.5) or when the efficacy of the vaccine was significantly lower among older populations; in these cases, prioritizing adults between 20-49 years resulted in the greatest reductions in mortality. A limitation of previous studies has been that they have mostly been based on age and individual connectivity has not been considered independently. While dividing the population in age groups is a practical and actionable solution for vaccine prioritization, it has the limitation of being a social construct with artificial cutoffs, which can sometimes mask the important role of the underlying contact structure.

Another challenge estimating the potential impact of COVID-19 preventive approaches has been that traditionally epidemiological models have depicted populations and epidemics as homogeneous [4], however the spread of COVID-19 has been characterized by a high variance in its reproductive number (R_t_) which leads to a pattern of super-connected individuals being responsible for most infections. [11, 12] This represents a major deviation from how influenza, which has a smaller R_t_ variance, spreads and requires adjustments to traditional models. The use of graphs to represent the contact structure of populations and the study of epidemics on networks have recently gained strong interest [13,14,15]. Hence, the aim of this study was to build on these approaches to qualitative estimate the role of individual connectivity and contact heterogeneity on the COVID-19 epidemic, and to identify the vaccination strategies that minimize infection and death based on the underlying contact structure of the population.

## Methods

We used a model that focuses on the interactions that infectious individuals have during the period when they are contagious. It is a middle-ground approach between agent-based [23] and mean-field [24,25,26] models, in the sense that we do not model every interaction that occurs in the population, as an agent-based model would, but we keep some randomness in the interactions instead relying on averages, which is one limitation of mean-field models. In line with the methodology previously applied by Holme (2017) [27], we consider a random graph of interactions, in which connections between nodes (i.e., individuals) represent interactions during the infectivity period (which we will call “risky interactions” or “risky contacts”).. In other words, we first choose at random the interactions individuals will have if they become infected. After setting up the underlying graph, we spread the infection on top in the form of a traditional SIR model, and let the infection spread like a contact process on top of the graph.

In order to assess how the contact structure of the population can affect the dynamics of an epidemics, we compared two types of populations, a homogeneous one, modelled by an Erdős-Rényi graph (ER) [16] and one with high variance in the number of contacts per individual, modelled by a graph with power-law degree distribution (PL) [17]. The degree distributions of both graphs are shown in Fig 1 and a visualization of their structure (with small N) is shown in S1 Fig. A detailed description of the construction of the graphs is presented next.

**Fig 1.**
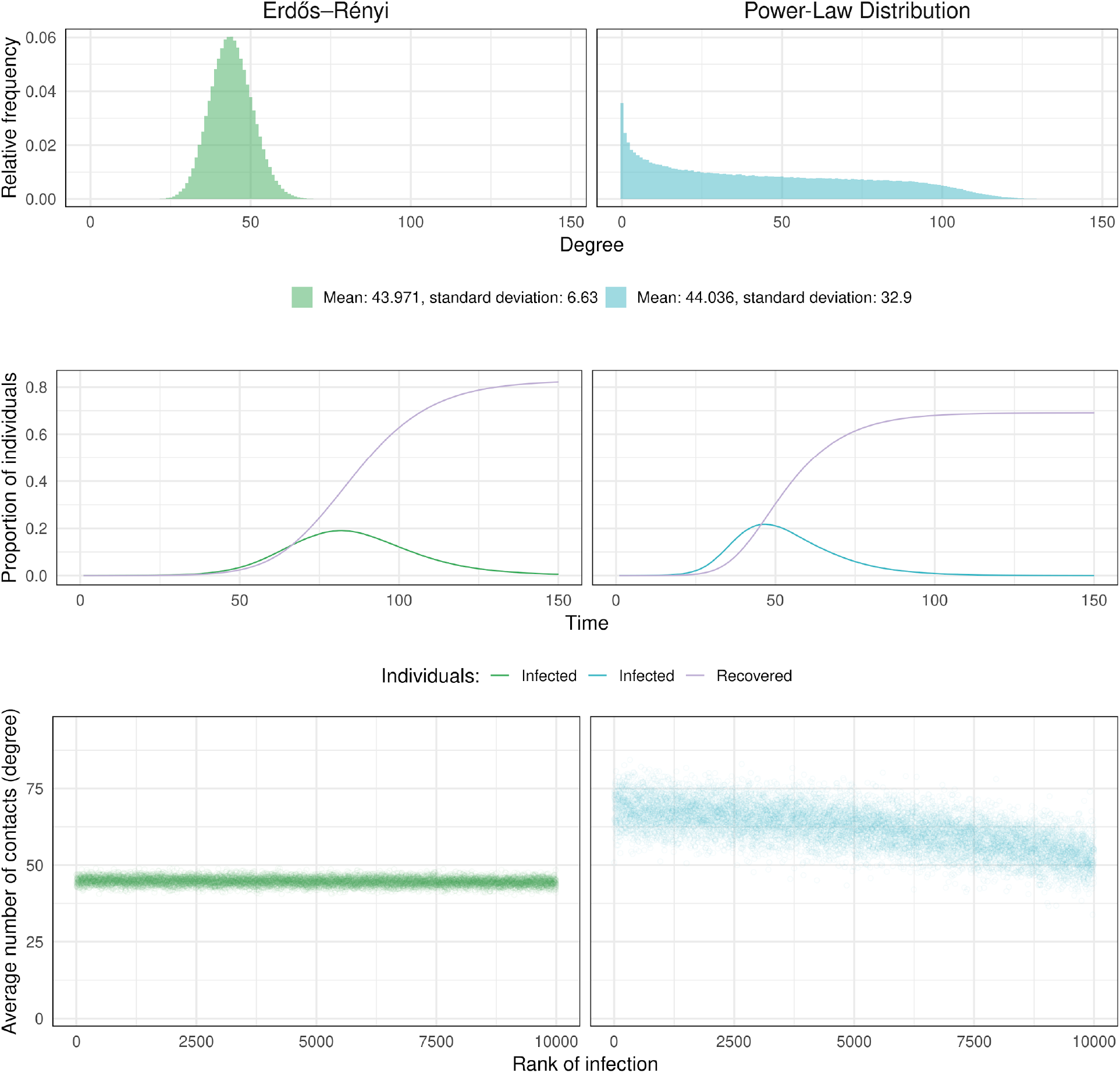
Two different ways of modelling social interactions. Top panels represent the distribution of the number of risky interactions in the ER and the PL graphs with 20000 individuals. Panels in the middle show a realization of the SIR process for each of the models. Bottom panels show the number of risky interactions individuals have, as a function of the order in which they are infected (dots show the average over 30 simulations).

### Interaction graphs

The Erdős-Rényi graph is one of the most classical random network models. [16] In a graph with *N* vertices, for any pair of vertices there exists an edge with probability 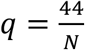, independently of each other. Hence, the number of connections per individual is binomially distributed with parameters *N* and *q*, which has mean *Nq* = *e* and variance *Nq*(1 – *q*).

To construct a graph with power-law degrees, we followed the method proposed by Qiao et al. Power-law [17] with exponent ***γ*** roughly means that the probability that a given vertex in the network has ***κ*** connections (in other words, it has degree ***κ***) behaves like ***κ***^**−*γ***^ for large ***κ*** if the total population size is large enough compared to ***κ***. [17] To construct this graph, to each individual ***i*** we assigned a random number ***δ***_***i***_, which is an exponentially distributed random variable of parameter ***λ*** (this parameter is related to the exponent ***γ***), and we fixed a positive parameter ***b*** (this parameter is related to the average number of connections per individual). Each pair of individuals (***i, j***) has risky interaction with probability 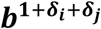. With this construction, we have 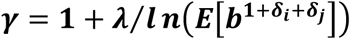, where ***E*** denotes expectation. In order to have e=44, we chose ***b*** such that 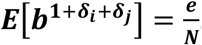 To do so, we used the moment generating function of an exponential random variable. We needed to choose ***b*** such that: 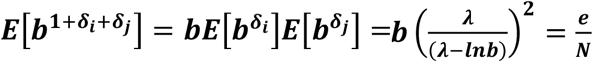 For this study, ***λ*** was set to 3, leading to ***γ***= 0.509.

### Infection dynamics

After simulating the structure of the interactions in the population, we simulated the spread of the infection using a SIR model, which is a continuous-time Markov chain where individuals can be in each of four states: Susceptible, Infected, Recovered or Dead. At time 0, we started with one infected individual chosen at random (the rest were all susceptible). Infected individuals could transmit the disease to susceptible individuals (if they had a risky interaction during the simulation). At the end of the infectious period, the infected individual either recovered or died. We assumed that recovered individuals could not be reinfected (S2 Fig, S1 Movie, S2 Movie).

In more detail, for each individual that gets infected (the ‘focal’ individual) the duration of the infectivity period is an exponential random variable with parameter ***T***_***r***_ = 10. This means that the expected duration of the infectivity period is 10 days. During this infectious period, the focal individual could each of their contacts with probability ***P***_***i***_. The way we implemented this in a Markovian way was by assigning to each of his neighbors an exponential random variable of parameter ***T***_***i***_. If one (or several) of these variables is smaller than the infectivity period, the focal individual transmitted the disease to the corresponding neighbor. All these exponential random variables were assumed to be independent. The parameter ***T***_***i***_ was chosen in such a way that the probability of infecting each of the susceptible neighbors ***P***_***i***_ was 0.05. To do so, we used a classical result on the minimum of two exponential random variables: 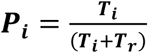.

At the end of the infectious period, the focal individual either died with some probability ***P***_***d***_ or recovered with probability **1** − ***P***_***d***_. The probability of death was initially set to 0.01 for all individuals and then modified based on connectivity for extended models.

The process was simulated using a Gillespie algorithm. Simulations were performed in R and code is available in a git repository (see Data Availability).

### Lockdown strategies

In our models, a lockdown corresponded to reducing the number of edges in the graph, i.e., decreasing the number of potential infectious contacts between individuals. We considered different lockdown strategies:

- Lockdown 1 (LD1): We removed each edge of the graph independently with some probability p = 0.5.
- Lockdown 2 (LD2): The number of contacts of any individual was bounded by M. To implement this strategy, we considered individuals sequentially. For each individual, if he has d contacts where d>M, we deleted M-d of these interactions at random. Otherwise, the number of his contacts remained unchanged.

In both cases, the lockdown started when the cumulative number of infected individuals reached a certain threshold (0, 10 or 30% of the total population) and it was lifted after a fixed number d of days. To provide a “fair” comparison, M was chosen in such a way that on average the same number of connections was removed in LD1 and LD2, in the Erdős-Rényi and power law cases. This corresponded to M= 26. However, in the power-law case, this choice yielded a very strong reduction of the number of edges (since most edges are carried by super-spreaders, that had a degree much higher than M). Therefore, in supplementary figures 4 and 5, we also show a variant of this strategy (Lockdown 2b), where M=82, which is the value needed to have a similar reduction in the number of edges as in LD1 for the power-law graph. This strategy has no effect in the Erdős-Rényi case since the variance in the number of contacts is smaller so it is very rare to find individuals with more than 82 connections.

### Vaccination strategies

In our model, vaccination corresponds to reducing the number of vertices with certain probability based on the effectiveness of the vaccine (efficacy *ϵ*. Each vaccination strategy corresponds to a way of subsampling *D* individuals in a population of size *N*. We implemented six different vaccination strategies that corresponded to different ways of choosing the sample of individuals to vaccinate.

- “Uniform”: D individuals uniformly selected at random for vaccination.
- “Most connected”: D individuals selected in order from most connected (those with the highest degree) to less connected.
- “Neighbor”: Randomly selected one individual and vaccinated a connection (i.e., one that is connected to the first one by an edge) also selected at random. This was repeated until D individuals were selected and vaccinated, ensuring that tendentially, individuals with a high number of connections got vaccinated. This is a real-life approximation following an approach that has been used to identify and vaccinate the most connected individuals, cf. [26].
- “Among most connected”: Subdivided the population into two groups of equal size: the “most connected” (those having the highest degree) and the “least connected” and chose D individuals uniformly at random among the most connected. This is a real-life approximation where groups which tend to be highly connected (i.e., healthcare workers, frontline workers) are prioritized for vaccination. A similar approach was suggested by Shahzamal et al. in the form of a vaccination strategy using an app to detect individuals that had been to high-risk locations. [28].
- “Among least connected”: Subdivided the population into two groups of equal size: “most connected” (those having the highest degree) and the “least connected” and chose individuals uniformly at random among the least connected.
- “Least connected”: D individuals selected in order from least connected (those with the lowest degree) to more connected.

We tested two variants of each of these strategies:

- Strategy S: selecting only among the susceptible individuals.
- Strategy SIR: selecting among the susceptible, infected or recovered (but not among the dead).

If vaccination is conducted at time 0, both strategies are equivalent because everyone is susceptible.

As a simplifying assumption, we considered that vaccinated individuals do not contribute to disease spread. It was assumed that in fully vaccinated individuals, the vaccine had an effectiveness of *0*.*9*. This means that among the *D* chosen individuals, only ϵD were effectively deleted from the graph. We tested three different vaccination starting times where vaccination started after the cumulative proportion of infected individuals reached 0%, 10% or 30%. Most models were conducted with vaccine doses available to vaccinate 25% of the population, but in an extended model we tested the impact of different interventions when the number of doses was doubled but the effectiveness (***ϵ***) was decreased from 0.9 to 0.5 to mimic the dose sparing strategies that have been applied in some countries.

As mentioned before, the probability of death was originally set uniformly to ***P***_***d***_ = 0.01, thus the number of infected is directly correlated with the number of dead. In extended models that assumed that old individuals were less connected and more vulnerable than younger individuals, we assigned a probability of death of ***P***_***d***_ = 0.07 to the 17% least connected and ***P***_***d***_ = 0.005 for the rest. This percentage corresponds to the proportion of individuals older than 65 in Utrecht and the probabilities are in line with the case fatality rate of COVID-19 for different age groups. [29, 30]

### Choice of the parameters

Since the simulations are computationally intensive and we wanted to test as many scenarios as possible, we chose a population size of 20000 (N). We also assessed if the sample size affected the results from the models with sample sizes varying from 5000 to 60000 (Fig S3). We observed that the fraction of infected individuals seemed to reach a stationary behavior for large N, which is a good indicator that, qualitatively, a larger population should behave similarly to our small world simulations.

The average number of risky interactions (e) was set to 44 following a study made in the city of Utrecht, Netherlands. [29] In this study, individuals of different age groups reported the number of people with whom they had a conversation of at least 10 minutes during a week, which we consider is a proxy for the number of risky interactions. We took the average of these numbers (according to the population census).

### Repeatability and sample size

The results presented consider independent realizations of the process. In a small number of simulations, the epidemic died out after infecting less than 0.5% of the population. These simulations were excluded from the results (because they are not consistent with the case of COVID-19). Results presented show the mean and standard error of 30 repetitions conditioned on infecting a macroscopic fraction of the population (>0.5%). Results are always expressed as the fraction of the population size N, instead of absolute numbers of individuals. For the purposes of this analysis, we considered interventions significantly different if over 95% of the simulations did not overlap.

### Key observables

In order to analyze the effect of connectivity of the underlying graph as well as the efficiency of lockdown and vaccination strategies, we consider different observables. These include the total number of infections after the end of the epidemic, the infection curve (which provides information about the maximum number of active cases as well as about the duration of the epidemic and the eventual emergence of secondary infection waves) and the mortality of the epidemic. Apart from these classical and commonly studied observables, we also consider the so-called rank-connectivity plots (cf. Fig 1 and Fig 4). These depict the degrees of the infected individuals in the order that they became infected. The shape and variance of this curve provides information about the relation between the connectivity properties of the graph and the dynamics of the epidemic, e.g., a decreasing curve corresponds to an epidemic where highly connected individuals tend to get infected first.

## Results

### Association of connectivity with time to infection

In the first model assessing the relationship between connectivity and time to infection, under the parameters described above (N=20000, e=44, *P*_*i*_=0.05), we computed the basic reproduction number R_0_ as the expected number of cases directly generated by one infected individual if all their neighbors are susceptible, i.e., R_0_ = e. *P*_*i*_ = 2.2. The effective reproduction number R_t_ (estimated from the simulations using 14 days rolling windows) varied from 1.7 at the beginning of the outbreak to 0.9 towards the end.

In baseline simulations, highly connected individuals tended to get infected early, while less connected individuals got infected at random times throughout the epidemic. This was especially clear for heterogeneous contact structures, where the average number of risky connections per individual at early stages was significantly higher and decreased with time (Fig 1). We also observed that the total number of people that were infected before the epidemics died out was smaller in the heterogeneous setting.

### Lockdown strategies based on contact structure

In a homogeneous population, Lockdown 1, where a restriction to mobility is imposed by the authority causing the removal of each interaction with some probability, and Lockdown 2, which imposed a maximum number of contacts per individual had similar effects, decreasing the overall number of infections from 0.84 ± 0.001 (without any intervention) to 0.77 ± 0.001 in both cases. The maximum number of infected individuals in a time period also decreased significatively. However, in the heterogeneous population Lockdown 1 had a greater effect minimizing the overall number of infections from 0.69 ± 0.001 (without any intervention) to 0.57 ± 0.001. However, Lockdown 2, which resulted in a smaller reduction in the overall number of infections (0.64 ± 0.001), had a significantly lower maximum number of infected individuals through time and there is a secondary wave of infections. In this case, Lockdown 2 is more likely to prevent a collapse of the health system, and thus could be a preferable strategy. Consistently with this, Lockdown 2 is the only one that has an impact in the average number of connections through time (Fig 2).

**Fig 2.**
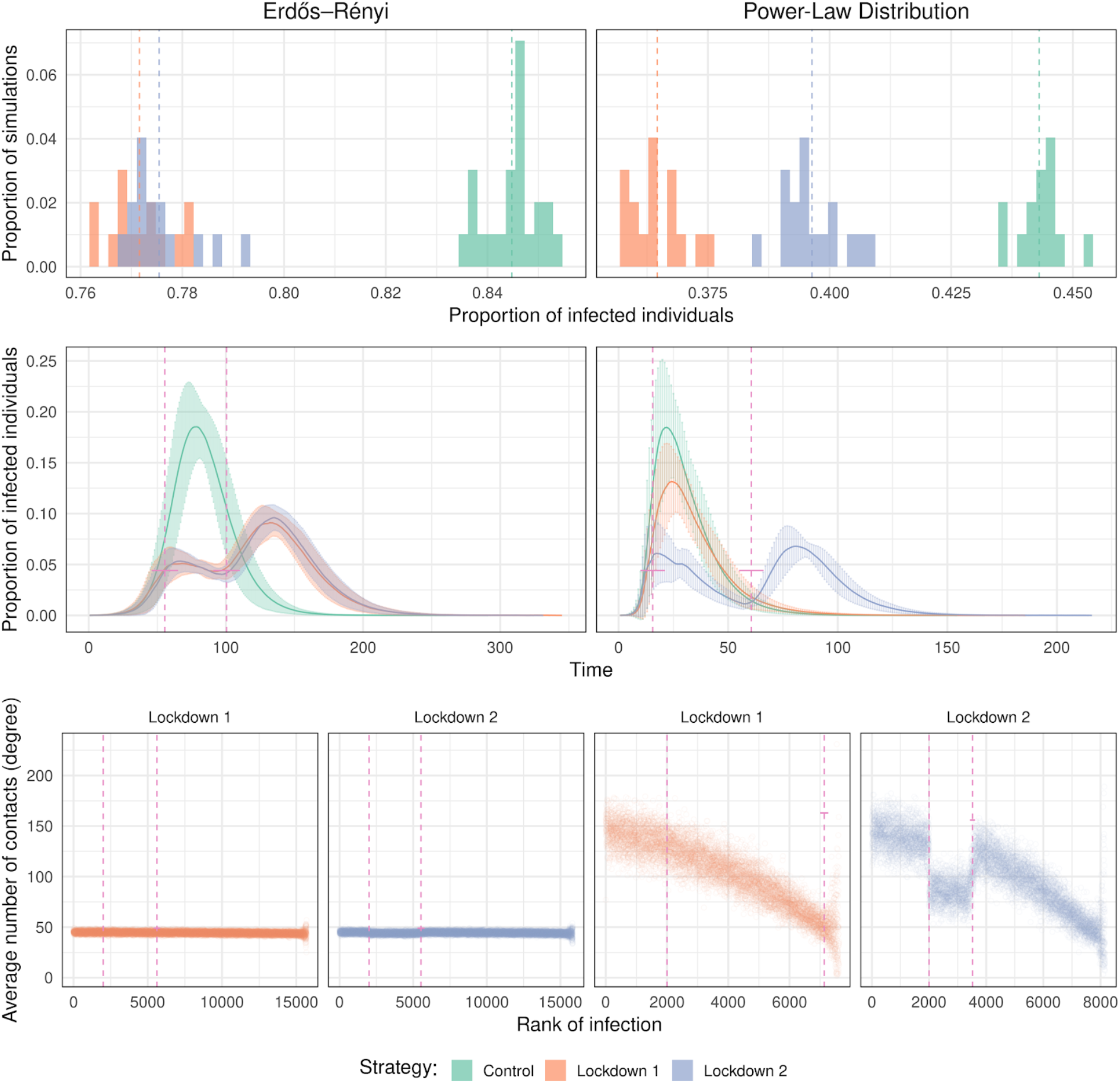
Effect of the different lockdown strategies. The lockdown starts when 10% of the population is infected and lasts for 45 days. Top panels represent the proportion of infected individuals at the end of the infection in 30 different simulations. The panels in the middle represent the number of infected people as a function of time. Error bars indicate standard deviation (computed from 30 repeats). Vertical bars indicate the start and the end of the lockdown. In the bottom panels, we show the average degree of the infected individuals as a function of their rank of infection for the two lockdown strategies. The controls are shown in Fig 1.

### Vaccination strategies based on contact structure

In the simulations with doses available for 25% of the population, vaccinating the most connected among the susceptible (S vaccination) resulted in the smallest proportion of infected individuals in homogeneous populations (0.45 ± 0.002 when vaccinating at time 0 vs 0.84 ± 0.001without intervention) although the effect was larger in the heterogeneous graph (0.18 ± 0.003 vs 0.69 ± 0.0007) (Fig 3). Similar results were obtained when varying the timing of the intervention. In models where susceptibility status was not considered (SIR vaccination), the benefits of targeting the most connected individuals decreased when more people had been infected before the intervention. For both graphs, vaccinating uniformly performed similarly to vaccinating the most connected when the intervention started after 30% of the population had already been infected. These strategies only reduced the proportion of infected from 0.84 ± 0.001 to 0.65 ± 0.003 in the homogeneous case and from 0.69± 0.0007 to 0.55 ± 0.002 in the heterogeneous case. (Fig 3). Vaccinating the least connected or among the least connected resulted in the highest proportion of infections in every scenario.

**Fig 3.**
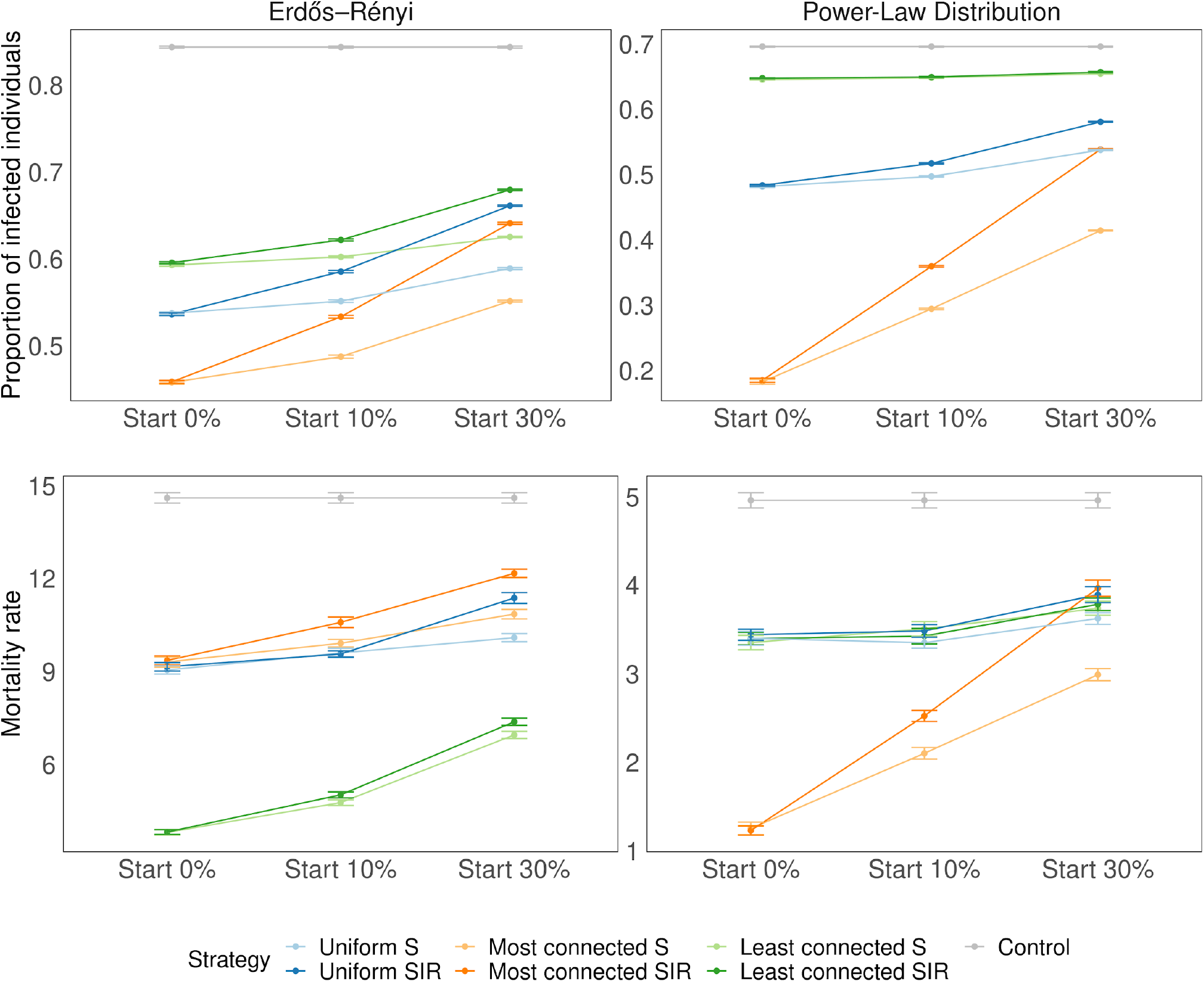
Proportion of infected and dead individuals for three vaccination strategies. The plot shows the proportion of infected at the end of the infection for 30 repetitions. The number of doses of the vaccine represents 25% of the population size (N = 20000). Error bars represent standard deviation. Different starting times are shown in the different panels (when 0, 10 and 30% of the individuals have been infected). On the top right panel, when vaccinating the most connected, the epidemic always died out quickly, before infecting at least 50 individuals, which is the minimum required to be considered a successful simulation (see Methods).

When the vaccine intervention was implemented early, when 10% of the population or less was infected, vaccinating the highly connected individuals first resulted in the greatest decline in the speed of propagation. However, when the intervention was implemented after 30% of the population had been infected, the decrease in the speed of propagation was much smaller (Fig 4).

**Fig 4.**
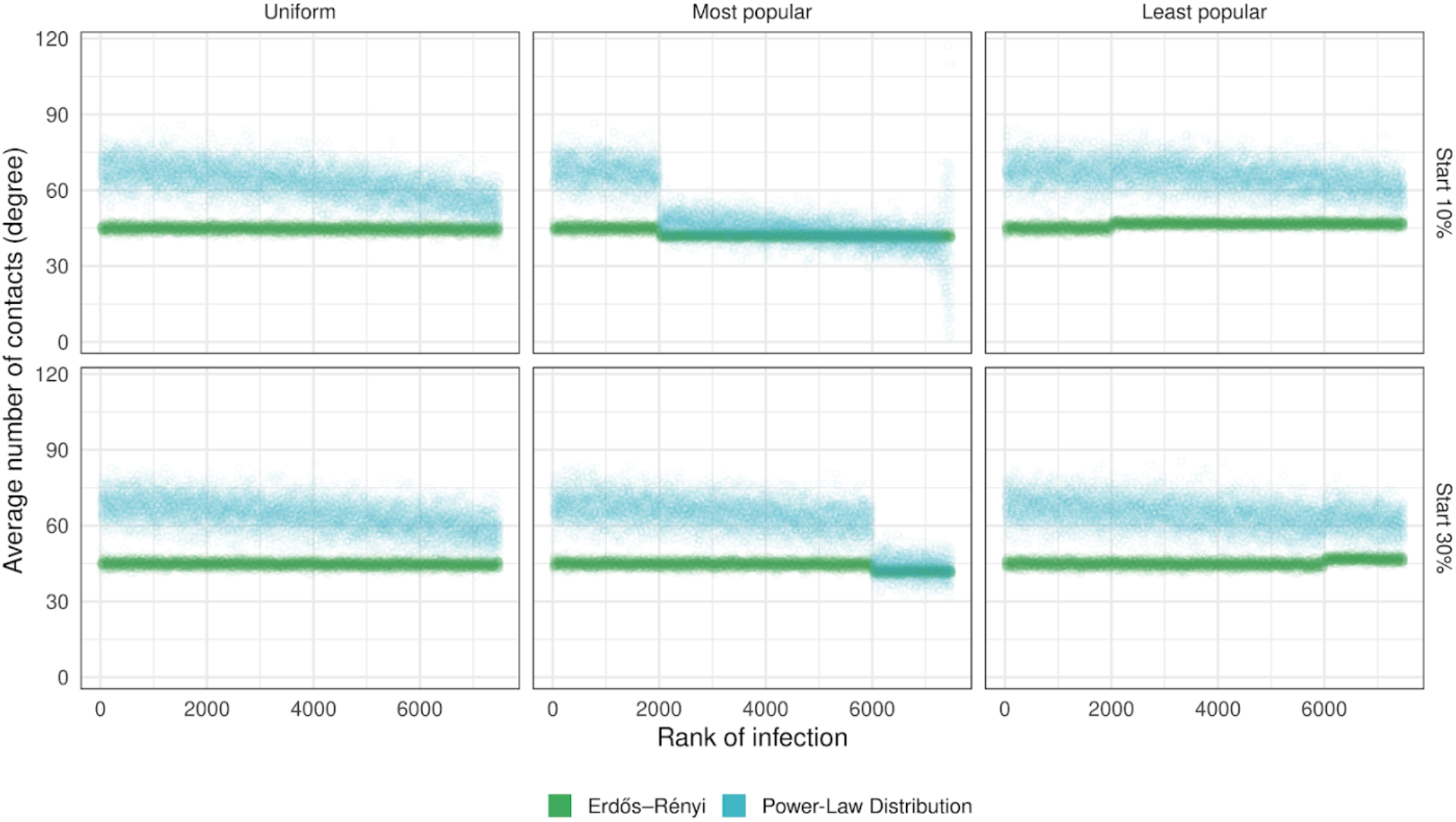
Effect of the vaccination strategy on the rank of infection. The dots represent averages over 30 simulations. The number of doses of the vaccine represents 25% of the population size (N= 20000). In these simulations we vaccinate individuals regardless of their status (S, I, R). Vertical bars indicate the time when the intervention is made.

### Vaccination strategies, connectivity, and mortality

In models where number of connections was linked to fatality rate, in the homogeneous case (ER), uniform and most connected strategies performed similarly, decreasing significantly the proportion of deaths from 0.014 ± 0.0001 in the control to 0.009± 0.0001 in both cases, when vaccinating at time 0. In this scenario, vaccinating the least connected (which were also the most vulnerable) was the best strategy yielding a proportion of deaths of only 0.004 ± 0.0001. Similar results were obtained when varying the time of the intervention. In a heterogeneous population (PL), the most connected (S and SIR) were the strategies that prevented more deaths, especially when the intervention started early, where the proportion of deaths was reduced from 0.004 ± 0.00008 in the control case to 0.0012 ± 0.00007. Uniform and least connected strategies performed equally well (0.003 ± 0.00007), preventing significantly less deaths than most connected S but slightly more deaths than the control. When vaccination started after 10 or 30% of the population had been infected, most connected S had an advantage over most connected SIR. (Fig 5).

**Fig 5.**
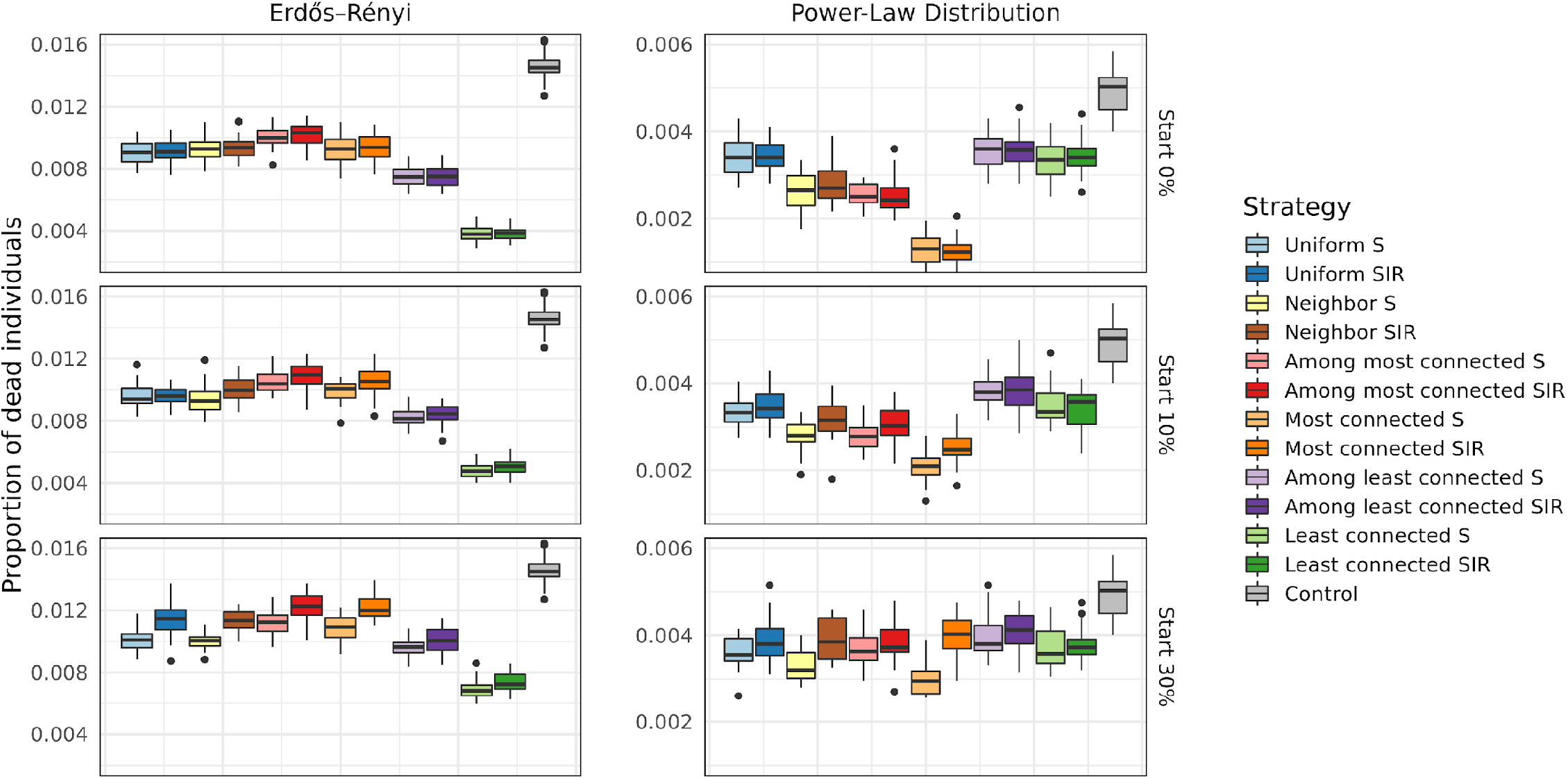
Proportion of deaths, for an effectiveness of 90% and for different values of *t*_*V*_ (top panels vaccination starts at 10% and bottom panels at 30%) The horizontal lines in the middle of the boxes show the mean values among all simulations, the upper and lower edges of the boxes are the quantiles q0.25and q0.75 corresponding to 75% respectively 25%. The vertical lines reach until q0.25-1.5*(q0.75-q0.25) downwards and until q0.75+1.5*(q0.75-q0.25). The points represent outliers (i.e., simulations whose results are atypical).

### Dose sparing under different vaccination strategies

In the case of a homogeneous population (ER), strategies that complete the vaccination schedule increasing effectiveness and those that vaccinated more people with lower effectiveness yielded similar results with respect to the total number of infected individuals. However, in the case of a heterogeneous population (PL), when vaccinating the most connected individuals, administering the two doses to fewer people was a more efficient strategy producing a smaller number of infected individuals. For the uniform strategy, the way of administering the doses made no difference. (Fig 6).

**Fig 6.**
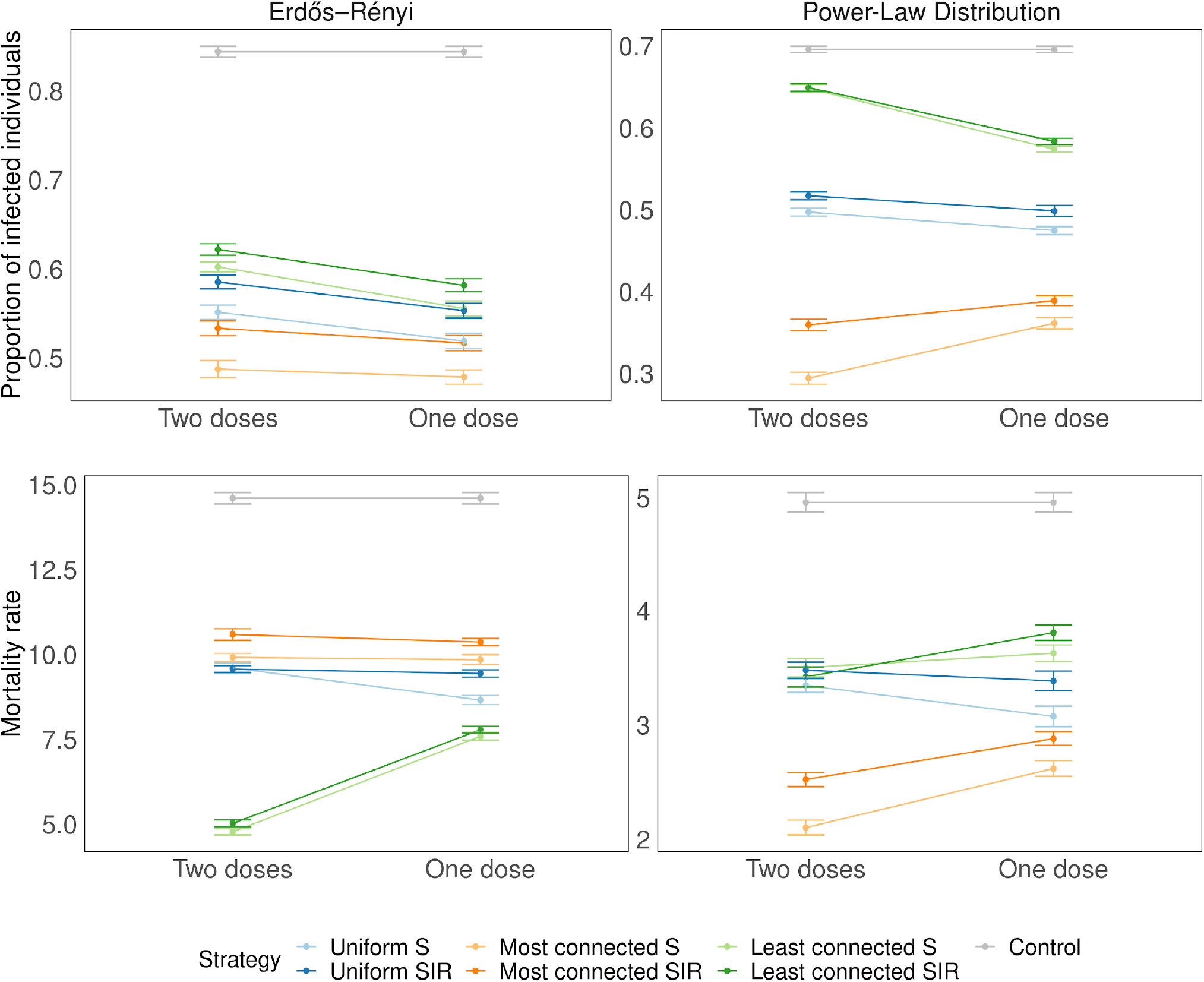
Dose sparing. Two doses: 25% of the population receives two doses of the vaccine (effectiveness 90%). One dose: 50% of the population receives a single dose of the vaccine (effectiveness 50%). Error bars represent standard deviation. The time of the vaccination *t*_*V*_ is when the cumulative number of infected individuals reaches 10%.

## Discussion

In this study, we modelled different lockdown and vaccination strategies based on connectivity. Our results confirm that heard immunity was reached earlier in heterogeneous scenarios, which is consistent with recent evidence from Britton et al. [4] Further, we also found that the level of heterogeneity in the underlying structure of risky contacts, also modified the effectiveness of different interventions such as lockdowns or vaccination approaches. This is particularly relevant for the case of COVID-19 where a few highly connected individuals have been responsible for most of the infections.

In order to account for differences in connectivity and heterogeneity, we used graph modeling approaches, which are gaining interest in the study of epidemics, [13,14,15] and developed an innovative application that allowed us to test preventive interventions on clearly distinct homogeneous and heterogeneous graphs. Previous studies had attempted to affect connectivity of graphs by removing uniformly chosen vertices from a random graph, [18,19,20] but found that, in the case of heterogeneous graphs to really affect connectivity, almost all vertices had to be removed, which limited their ability to test interventions. In an alternative approach, [21], the authors studied the propagation of an epidemic (SIS) on a heterogeneous graph when removing some special vertices, chosen using fine properties on the underlying graph and interpreted it as a prioritization strategy. Our approach, using two distinct fixed underlying graphs and considering different strategies to remove vertices, allows us to make fair comparisons of different prioritization strategies under different context. Moreover, this approach opens the possibility of testing interventions at different times of the epidemic. Furthermore, the models presented in this paper, do not require perfect knowledge of the underlying contact structure to generate valuable recommendations, this is particularly important for public health because usually this information is also unknown to decision makers fighting an epidemic.

We first used our models to assess the role of heterogeneity in the case of lockdown interventions. We compared two strategies: Lockdown 1 which can be interpreted as the result of a general stay-home initiative and Lockdown 2 which could correspond to protective measures like the prohibition of events with many participants or restriction of the number of individuals who are allowed to meet at the same time. We demonstrated that the best intervention depends on the underlying contact structure of the population. In a homogeneous population, both strategies are equivalent, while in the heterogeneous population Lockdown 2 is more likely to prevent a collapse of the health system, and thus would be a preferable public health strategy.

We then used our model to assess the efficacy of different vaccination strategies. In the heterogeneous context, interventions that prioritize more connected individuals performed better at preventing infection and deaths when compared with uniform strategies where vaccines are distributed at random. Conversely, strategies that prioritized the less connected individuals had the worst outcomes even when a higher fatality rate was assigned to the less connected. Most current COVID-19 strategies begin vaccination with highly connected individuals (healthcare workers, populations living in elderly care facilities) but then turn to vaccinate older and vulnerable populations which tend to be among the least connected [31]. If in fact more vulnerable individuals are less connected, current vaccination strategies that prioritize older and more vulnerable individuals over younger and more connected might be suboptimal to address the COVID-19 pandemic.

Another important result is the importance of intervention timing during the history of the epidemic. Especially under heterogeneous contact structures that are similar to the COVID-19 spread pattern, highly connected individuals tend to get infected very early and drive the early stages of the contingency. This is consistent with what was reported by Hoffmann et al. that also found that in a heterogeneous graph, the highly connected individuals are over-represented in the group of infected individuals at the initial phase of an epidemic. [22] Besides studying this finding in relationship with lockdown strategies, we also expanded it to study the role of timing on the impact of vaccination strategies. Our results connect this radical contact-driven transformation of the topology of the graph over time with the success or failure of intervention targeting most connected individuals depending on the time of the intervention. Further, this finding highlights the importance of considering what proportion of the population has already been exposed to the virus and potentially developed some immunity when the intervention is implemented. We observed that in the absence of susceptibility-based targeting, interventions prioritizing highly connected individuals were more effective early in the epidemic. Others have highlighted the importance of antibody testing to prevent infection and death, [10, 32] and our results further support this approach especially when targeting highly connected individuals late in an epidemic when a high proportion has already been infected. The proportion of healthcare workers with SARS-CoV-2 antibodies ranges between 2-50% in different settings, [33] based on our results, targeting the available doses based on previous immunity could maximize its impact especially in settings where seroprevalence is high.

A challenge of these connectivity-based strategies is that identifying the most connected individuals in real life is not easy. To address this, we tested two modified versions of the “most connected” vaccination strategy (see S6 Fig and S7 Fig). The first one consists of randomly selecting an individual and then vaccinating a person connected to them (*neighbor*), which biases the selection towards the most connected. For the second one, we divided the population into two groups based on their number of connections (*most and least connected*) and then sampled among the most or least connected group. In the heterogeneous scenario, vaccinating the most connected remained the most effective approach; however, *neighbor* and *among the most connected* also performed better than *uniform*, supporting the targeting of highly connected individuals as a promising strategy for COVID-19 vaccination. An operationalization of vaccinating among the most connected would be to prioritize individuals with occupations that require face-to-face interactions, for example those in the service industry.

A criticism of targeting the most connected individuals has been that these tend to be young and less vulnerable [31], hence we also extended these models to assess their impact on mortality under the assumption that connectivity is inversely associated with fatality. In the homogeneous structure (ER), vaccinating the most vulnerable resulted in reduction in the number of deaths. However, in heterogeneous structures, strategies targeting more connected individuals performed better than *uniform* and *least connected*, especially when targeting the susceptible. Uniform approaches have the advantage of being easier to implement and they perform significantly better than targeting less connected individuals, which resulted in the greatest number of infected. According to our models, the uniform approach is not always bad, vaccinating just 25% of the population with this strategy reduced infections compared to the control and, in some cases of late intervention, it can even outperform vaccinating the more connected individuals. Some counties in the US started implementing uniform approaches because they originally faced challenges distributing vaccines to older populations first, [34] our results support this vaccination approach or those prioritizing frontline workers over targeting less connected individuals.

Most of the vaccines that are available for COVID-19 were designed to be administered in two doses (and the effectiveness was shown to be over 90%). However, it has been suggested that governments should aim to give as many people as possible a single dose, instead of using half the vaccines currently available on second doses (i.e., dose sparing). [35] The effectiveness after a single dose has been estimated to be around 50%. In the case of a heterogeneous population, when vaccinating the most connected individuals, administering the two doses was a more effective strategy producing a smaller number of infected individuals. However, when the least connected were prioritized, dose sparing resulted in fewer infections, suggesting that if current strategies that prioritize less connected individuals continue, applying a single dose to more people would be the best approach (although still significantly worse than targeting the most connected or even distributing the vaccine at random).

A strength of this study is the middle-ground modelling approach between agent-based [21] and mean-field [22] models, that combines the dynamic nature of the first with the computational efficiency of the latter. This is achieved by focusing solely on the interactions that infectious individuals have during the period when they are infectious, a crucial part of the social dynamics of the propagation of the virus. While our method disregards some features of a real society, it allows us to capture the essential differences between homogeneous and heterogeneous contact structures. In this sense, a limitation of these models is that they do not provide quantitative estimates of the exact impact of different interventions, however they provide sound qualitative judgments that allow to rank different vaccination strategies based on number of infections and deaths. Similarly, the graphs used in this study were parameterized based on data from a single study from a European city and have a relatively small sample size compared to most urban areas, however modifying the average number of contacts or the sample size did not modify our findings.

In conclusion, the effectiveness of vaccination strategies depends on the heterogeneity of the underlying contact structure and the timing of the intervention, it is important to consider this when implementing COVID-19 preventive approaches. Future applications of these models that were specifically designed to include differences in heterogenicity include the study of COVID-19 adaptation in these different scenarios and applications to other preventive approaches or infectious diseases.

## Data Availability

Code is available at our github repository.

https://github.com/JulioNava31/The-role-of-connectivity-on-COVID-19-preventive-approaches.

## Data availability

Code is available at: https://github.com/JulioNava31/The-role-of-connectivity-on-COVID-19-preventive-approaches.

## Acknowledgements

We would like to thank Arno Siri-Jégousse, Fabian Freund, Ximena Escalera-Fanjul, Karla Galaviz, Gloria Soberón, Adithya Cattamanchi, April Mohanty, Anarina Murillo and Bernardo Flores for fruitful discussions.

## Author contributions

VMP: Investigation, Methodology, Formal Analysis, Validation, Supervision, Writing-Review & Editing; JNT: Formal analysis, Software, Visualization. AT: Methodology, Investigation, Writing-Review & Editing, EN: Software, Formal Analysis, Investigation; ACGS: Conceptualization, Methodology, Supervision, Writing – Review & Editing; IGC: Writing – Original Draft, Resources, Supervision. All authors analyzed the results of the simulations and approved the final manuscript.

## Supplementary Figures

**S1 Fig.**
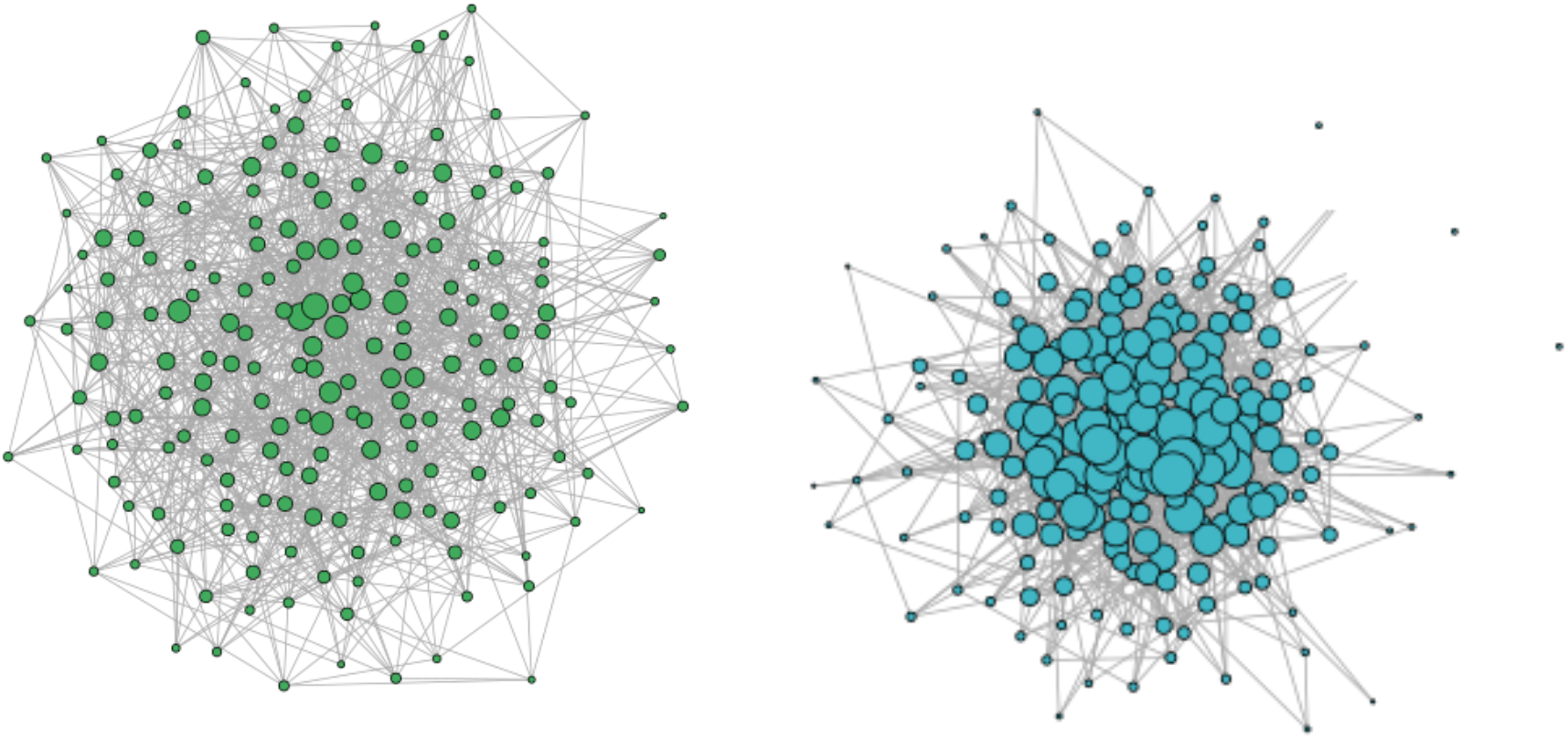
An Erdős-Rényi graph (left hand side) and a power-law degrees graph (right hand side). For an easy visualisation, the parameters where set to N = 200, e = 12, *λ*=3.

**S2 Fig.**
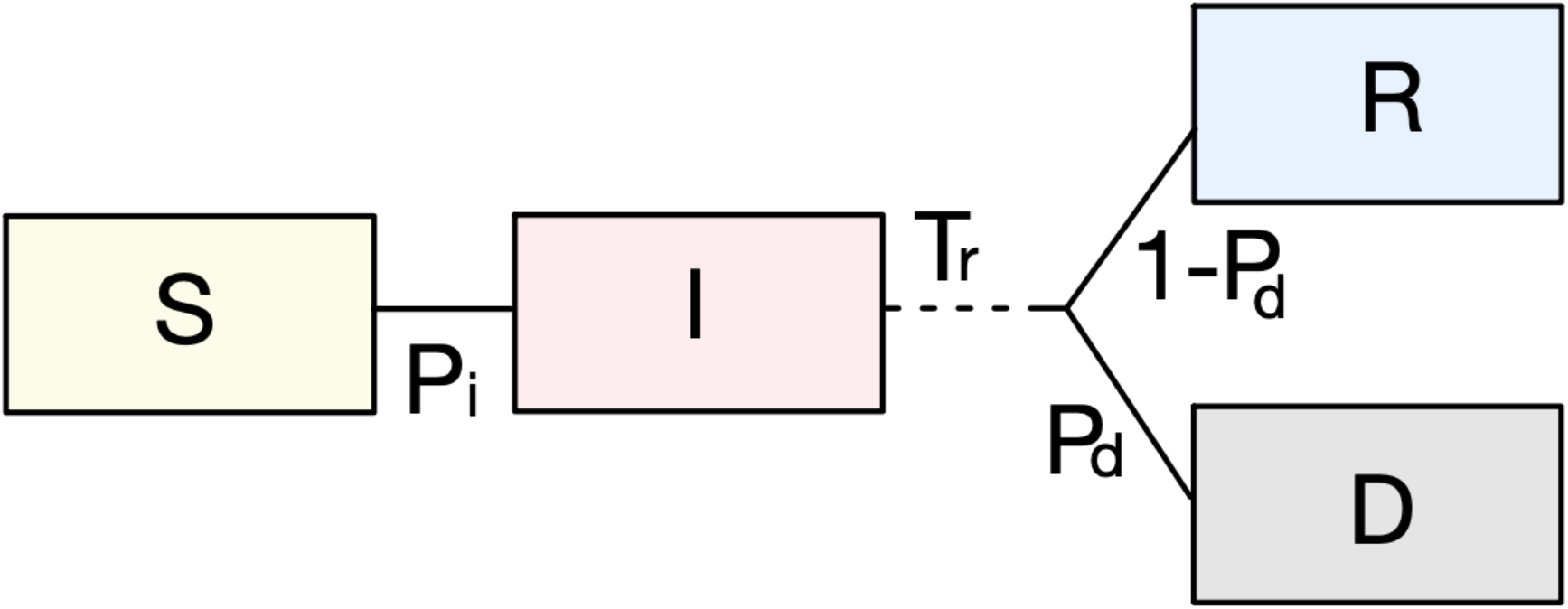
Transition rates of our SIR model. Susceptible individuals can become infected with probability *P*_*i*_ if they have an infected neighbour. Infected individuals remain infected for an exponential random time with mean *T*_*r*_. At the end of this infectious period they can recover with probability 1 – *P*_*d*_ or die with probability *P*_*d*_.

**S3 Fig.**
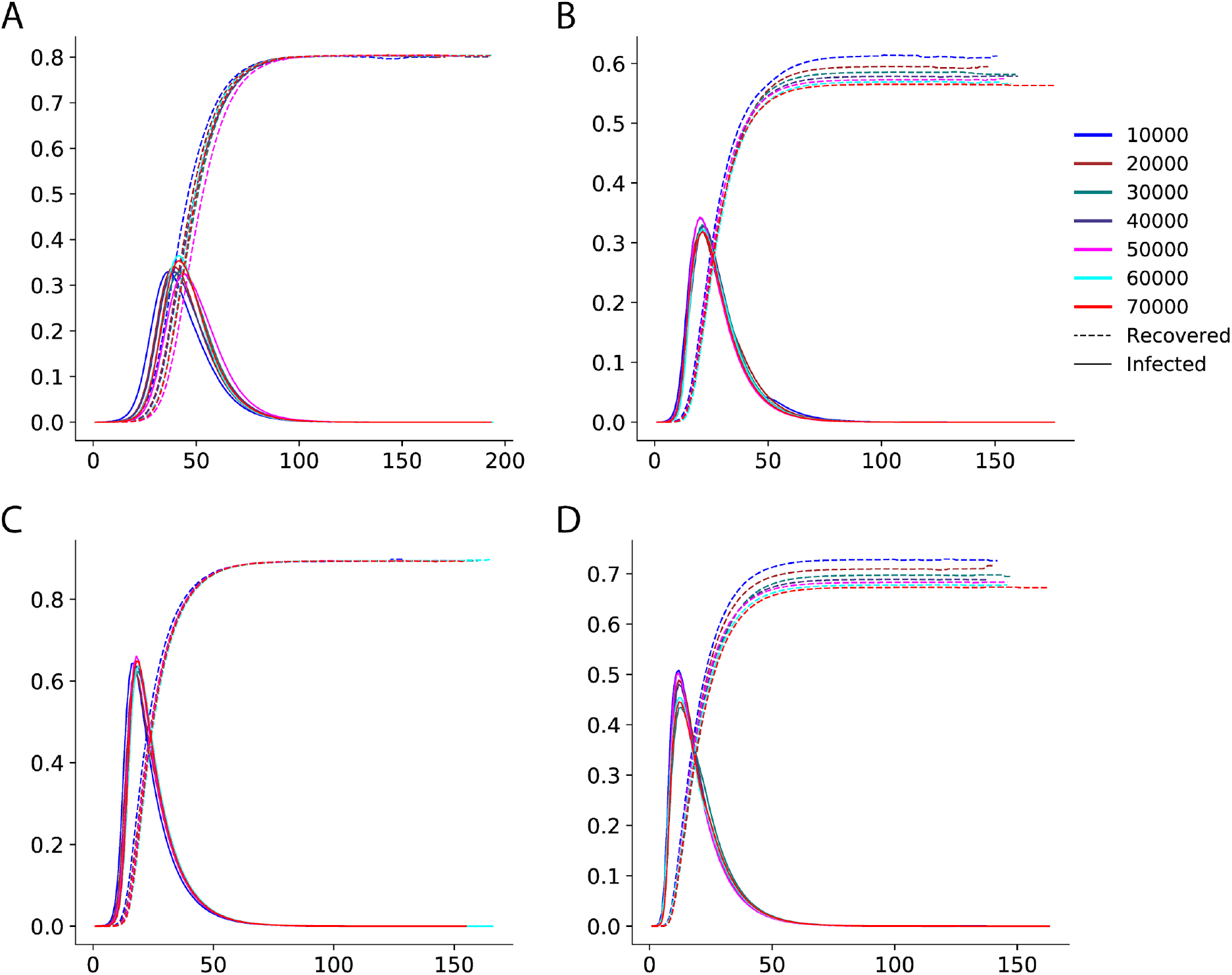
Infected and recovered curves for population sizes 10 000, 20 000, 30 000, 40 000, 50 000, 60 000 and 70 000. The curves represent the proportion of individuals in each category as functions of time. Each curve corresponds to the mean over 30 repetitions. A, C: Erdős-Rényi graphs, B, D: Power-law degree, A, B: e = 5. C, D: e = 10. The rest of the parameters are *p*_*i*_ = 0.5 and *λ*=3.

**S4 Fig.**
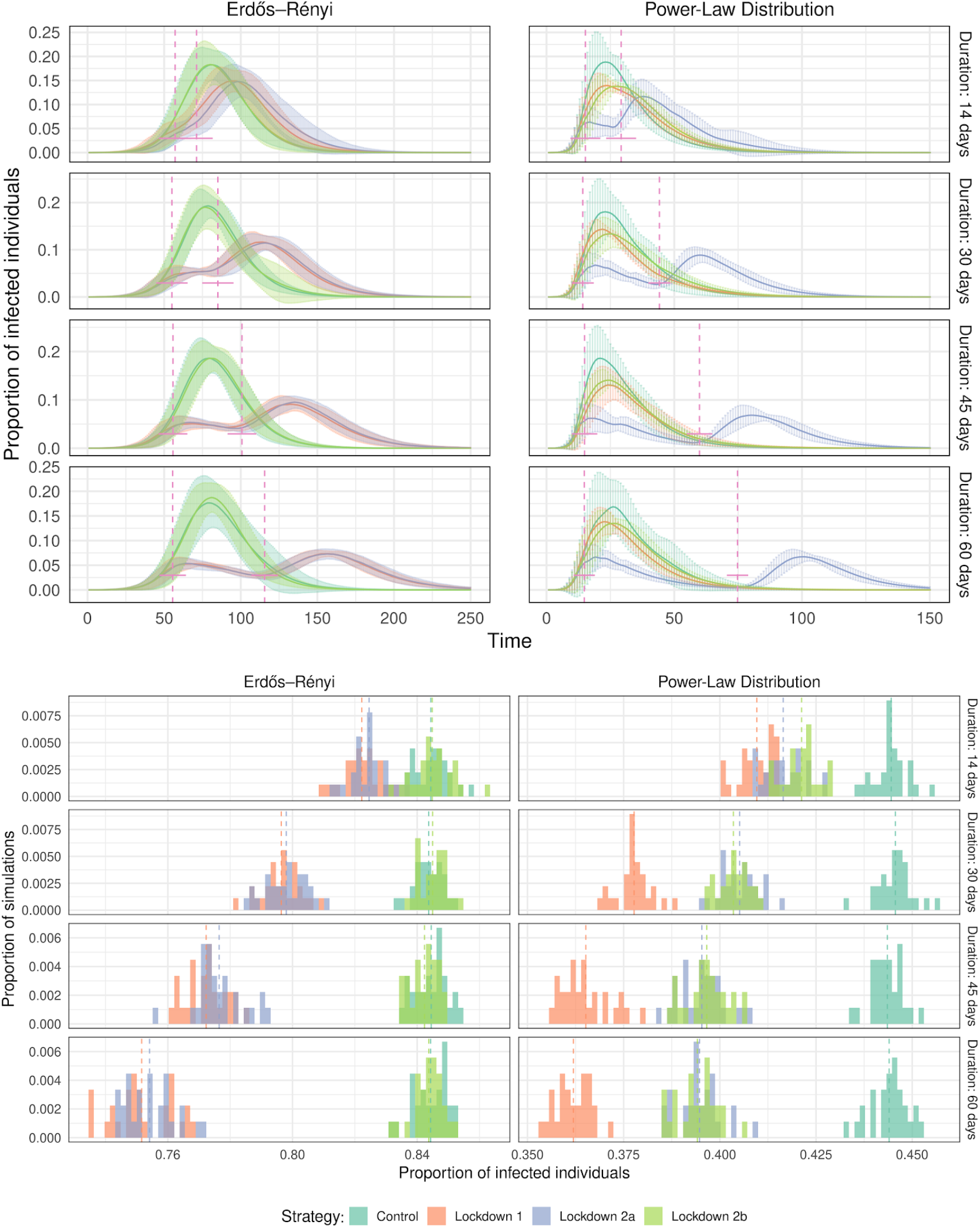
Effect of the duration of the lockdown for different strategies. Top panels: proportion of infected individuals through time. Bottom panels: distribution of the total number of infected individuals for 30 different simulations. The lockdown starts when the cumulative number of infected individuals is 10%. In the top panels, the dashed lines show the beginning and the end of the lockdowns (and its standard deviation indicated by a horizontal line). In the bottom panel dashed lines correspond to the average proportion of infected individuals for each condition.

**S5 Fig.**
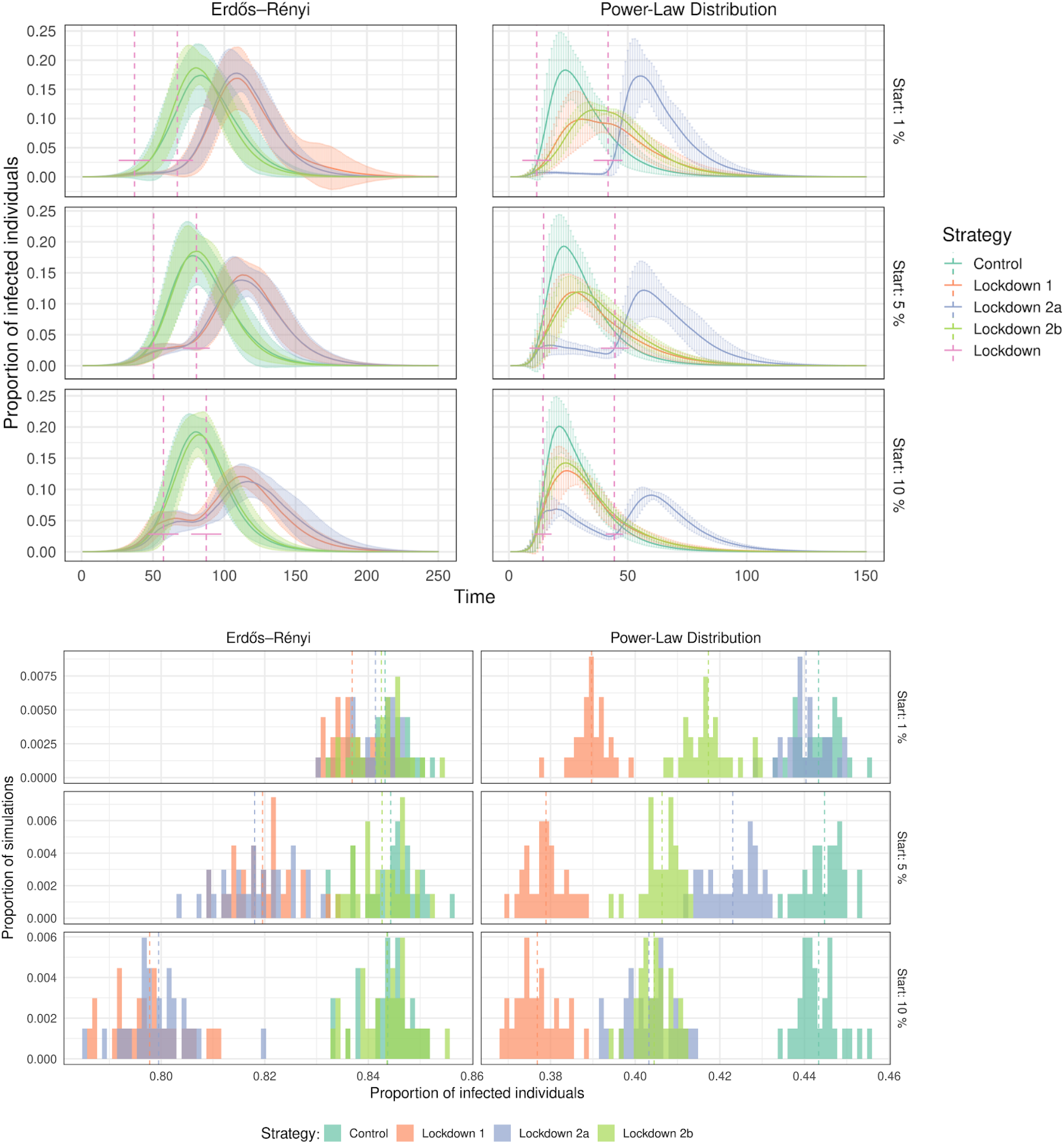
Effect of the starting time for the lockdown. The duration is fixed as 30 days, and we vary the cumulative proportion of infected individuals at the start of the lockdown tL. Top panels: proportion of infected individuals through time. Bottom panels: distribution of the total number of infected individuals for 30 different simulations. The dashed lines have the same meaning as in S3 Fig. Observe that for tL=1%, lockdowns have no substantial effect on the maximum of the infection curve in the Erdős-Rényi case, and the same holds for lockdown strategy 2a in the power-law degree case. A lockdown started later (at tL=5% or 10%) is better with this respect.

**S6 Fig.**
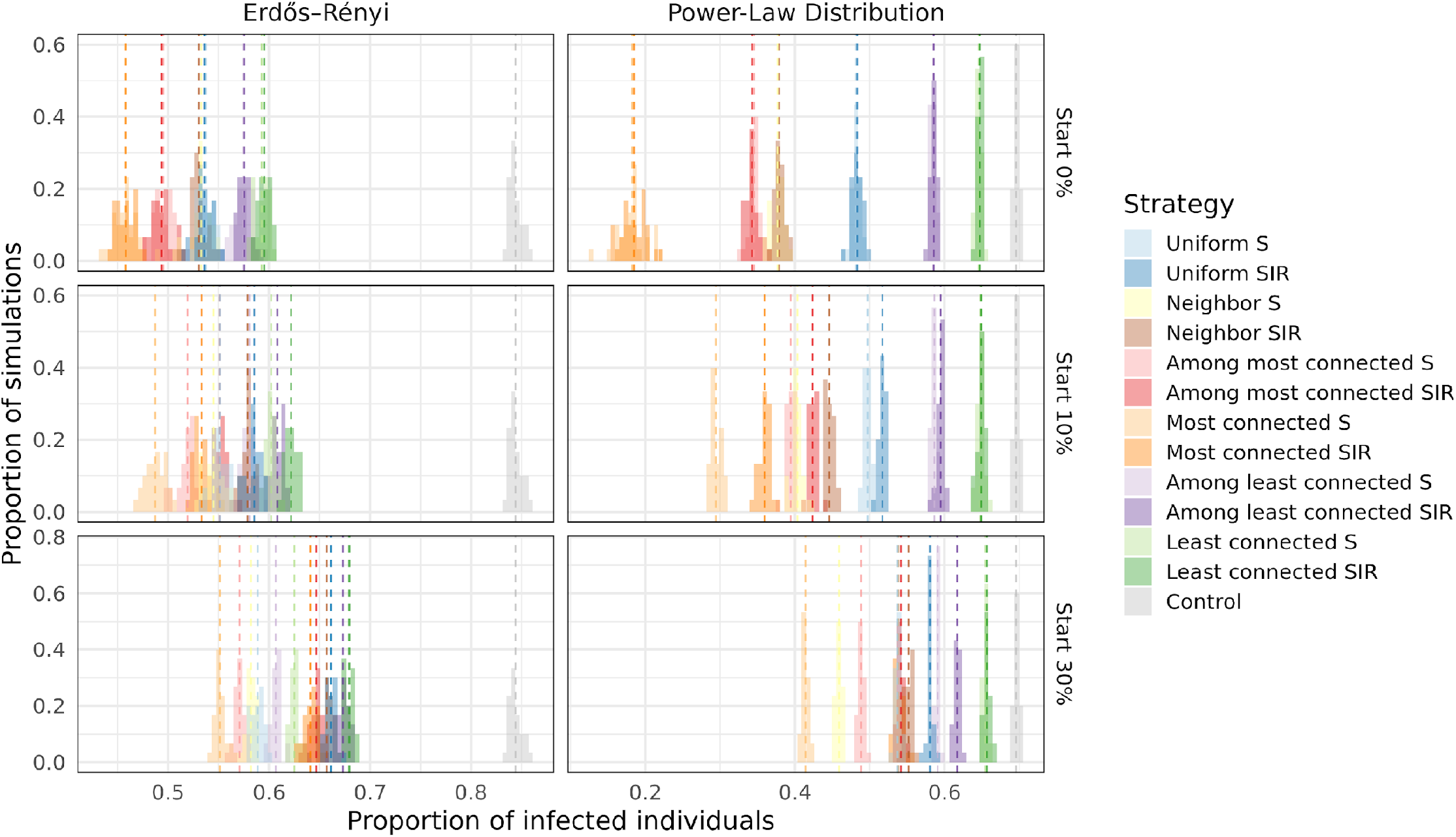
Proportion of infected and dead individuals for all the vaccination strategies. The plot shows the proportion of infected at the end of the infection for 30 repetitions. The number of doses of the vaccine represents 25% of the population size (N = 20000). Different starting times are shown in the different panels (when 0, 10 and 30% of the individuals have been infected). On the top right panel, when vaccinating the most connected, the epidemic always died out quickly, before infecting at least 50 individuals, which is the minimum required to be considered a successful simulation (see Methods).

**S7 Fig.**
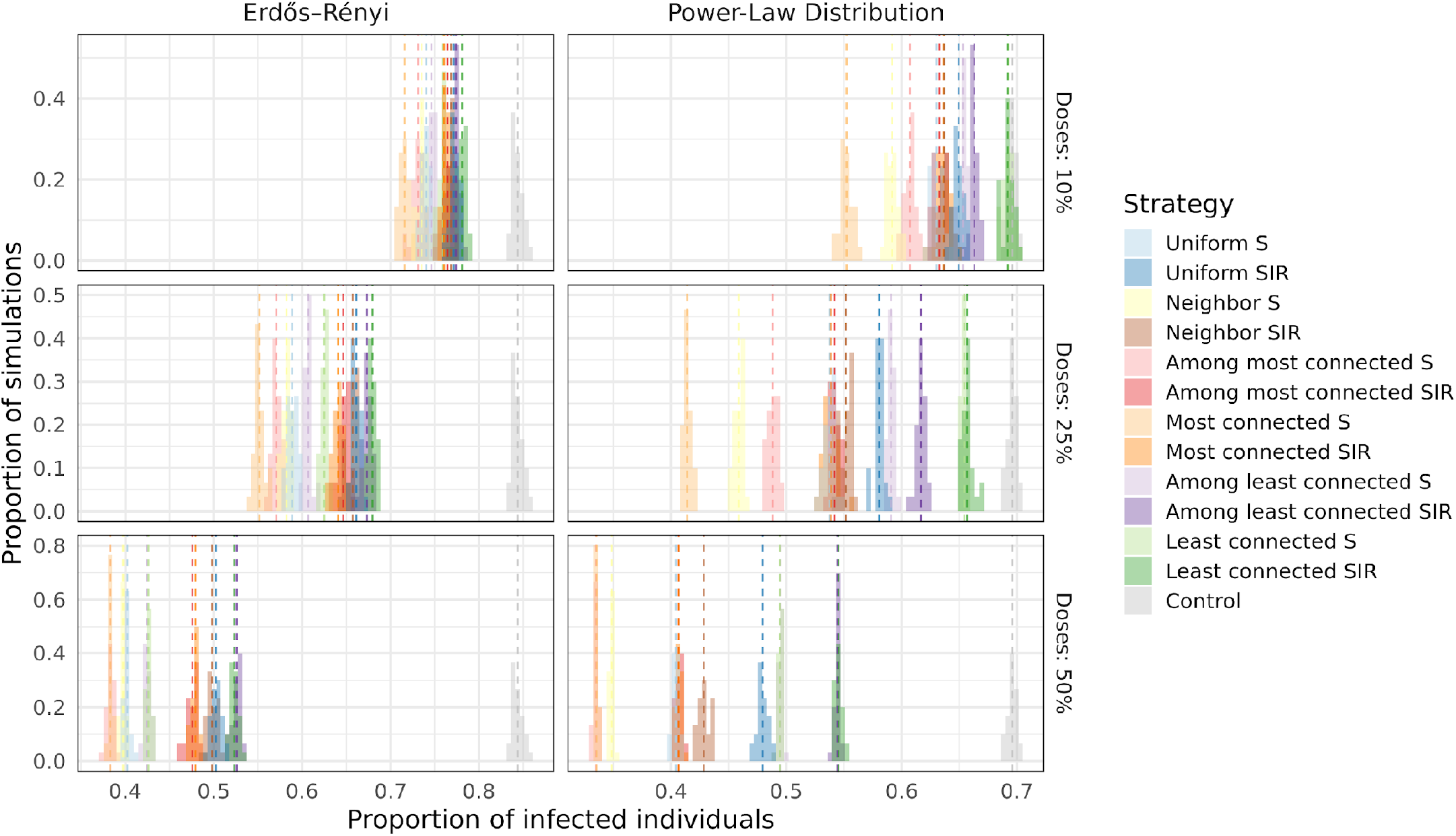
Effect of the number of doses of the vaccine. Plots show the total number of infected individuals in 30 simulations for the Erdős-Rényi (right) and power-law (left) graphs. From top to bottom we increase the number of individuals that we can vaccinate (10, 25, 50%). The time of vaccination is when the cumulative number of infected reaches 30% of the populations (*t*_*V*_ = 30%).

## Supplementary Materials

**S1 Movie**.

Propagation of the epidemics in an Erdős-Rényi graph. Vertices are colored depending on the status of the individual they represent. Blue: susceptible, red: infected, green: recovered, black: dead. The edge connecting *i* and *j* is colored in red when individual *i* infects individual *j*. The numbers that appear at the end are the number of individuals that were infected by each individual.

**S2 Movie**.

Propagation of the epidemics in a power-law degree distribution graph. Vertices are colored depending on the status of the individual they represent. Blue: susceptible, red: infected, green: recovered, black: dead. The edge connecting *i* and *j* is colored in red when individual *i* infects individual *j*. The numbers that appear at the end are the number of individuals that were infected by each individual.

